# CARE: a novel wearable-derived feature linking circadian amplitude to human cognitive functions

**DOI:** 10.1101/2023.04.06.23288232

**Authors:** Shuya Cui, Qingmin Lin, Yuanyuan Gui, Yunting Zhang, Hui Lu, Hongyu Zhao, Xiaolei Wang, Xinyue Li, Fan Jiang

## Abstract

Circadian rhythms play a critical role in regulating physiological and behavioral processes, with amplitude being a key parameter for their characterization. However, accurately quantifying circadian amplitude in natural settings remains a challenge, as traditional melatonin methods require lab settings and are often costly and time-consuming. Wearable devices are a promising alternative as they can collect consecutive 24-h data for multiple days. The most commonly used measure of circadian amplitude from wearable device data, relative amplitude, is subject to the masking effect of behaviors and fails to leverage the rich information in high-dimensional data, as it only uses the sum of activity counts in time windows of pre-specified lengths. Therefore, in this study, we firstly proposed a pipeline to derive a novel feature to characterize circadian amplitude, named **c**ircadian **a**ctivity **r**hythm **e**nergy (CARE), which can well address the above-mentioned challenges by decomposing raw accelerometer time series data, and then we validated the new feature CARE by assessing its correlation with melatonin amplitude (Pearson’s *r* = 0.46, *P =* 0.007) in a dataset of 33 healthy participants. Secondly, we investigated its association with cognitive functions in two datasets: an adolescent dataset (Chinese SCHEDULE-A, n = 1,703) and an adult dataset (the UK Biobank dataset, n = 92,202), and we found that the CARE was significantly associated with the Global Executive Composite (*β* = 28.02, *P* = 0.016) in adolescents, and reasoning ability (OR = 0.01, *P <* 0.001), short-term memory (OR = 3.42, *P <* 0.001), and prospective memory (OR = 11.47, *P <* 0.001) in adults. And finally, we explored the causal relationship using Mendelian randomization analysis in the adult dataset. We identified one genetic locus with 126 SNPs associated with CARE using genome-wide association study (GWAS), of which 109 variants were used as instrumental variables to conduct causal analysis. The results suggested that CARE had a significant causal effect on reasoning ability (*β* = -59.91, *P* < 0.0001), short-term memory (*β* = 7.94, *P* < 0.0001), and prospective memory (*β* = 16.85, *P* < 0.0001). The findings suggested that CARE is an effective wearable-based metric of circadian amplitude with a strong genetic basis and clinical significance, and its adoption can facilitate future circadian studies and potential interventions to improve circadian rhythms and cognitive functions.

## Introduction

Circadian rhythms synchronized by the rotation of the Earth are 24-h self-sustained oscillations that can influence a range of physiological responses and behavioral processes, such as hormone secretion and sleep-wake cycles[1–5]. The circadian rhythms are regulated by a complex transcriptional feedback loop involving the “core circadian clock” in the suprachiasmatic nucleus (SCN) of the hypothalamus and the other peripheral clocks throughout the body[6, 7]. Rhythmic fluctuations driven by the core clock could be illustrated as a sinusoidal curve with a period of about 24 hours[8], and hence the circadian rhythms are typically described using three key parameters (**Supplementary Figure 1**): period (the length of one cycle), phase (the timing of a reference point in the cycle relative to a fixed event), and amplitude (the difference between crest and trough values of the cycle)[9]. While methods for measuring period[10, 11] and phase[12–17] are well-established and accurate in both laboratory and field studies, assessing circadian amplitude remains challenging.

Although there is no accepted gold standard for measuring the circadian amplitude[18], the amplitude of pineal hormone melatonin has been used as one of its reliable biomarkers, and changes in the melatonin amplitude are also observed in forced desynchrony studies [19, 20]. However, assessing the melatonin amplitude requires a melatonin profile of at least 24 hours with continuous collection of blood or saliva samples[21]. Given that the collection protocol of melatonin is costly, labor-intensive, and infeasible for large population studies and intensive follow-up cohort studies in natural settings, researchers have attempted to find alternative methods to characterize circadian amplitude using wearable sensor data[22, 23].

Wearable devices can continuously monitor the physical activity of an individual for multiple days in a natural environment, and thus can solve the challenges present in melatonin sampling. The main rationale is based on the biological knowledge that both the melatonin rhythms and rest-activity cycles are strongly influenced by the core circadian clock and hence they may share similar oscillation patterns[8]. Nevertheless, accelerometer data also have some drawbacks. A major challenge is that the wearable device-derived circadian amplitude is influenced by voluntary behavioral changes in the natural environment that are not related to circadian rhythms[12, 17, 18]. More importantly, the most commonly used accelerometer-derived measure, relative amplitude, is calculated solely based on the differences between the highest and lowest activity counts[22, 24], which has lost rich information of the high-dimensional activity data. Furthermore, the direct relationship between the circadian amplitude characterized by accelerometer data and the circadian amplitude depicted by melatonin data has not been well studied. Thus, it is critical to establish and validate a sound method for extracting accurate information about circadian amplitude from high-dimensional accelerometer data to fully leverage the rich high-dimensional information without the masking effect of behaviors.

Changes in the amplitude of melatonin have been widely observed in age-related cognitive impairments and neurodegenerative diseases[9, 25–28]. However, few studies investigated the association between circadian amplitude and cognitive function in general population. Given that the circadian rhythms vary greatly during adolescence and many mental disorders also first appear in the adolescent stage[29, 30], and that the circadian rhythms are susceptible to heavy life and work during adulthood, it is necessary to examine the correlation between circadian amplitude and cognitive performance at these stages of life[31]. Furthermore, currently there is no evidence to support a causal relationship between circadian amplitude and cognitive performance[9, 32]. One prior study tried to study the causal relationship, but it only measured the relative amplitude and found that it was only associated with a small number of genetic variants that could not be used to investigate the causal relationship using Mendelian randomization (MR) analysis[33]. Thus, it is necessary to develop a metric of circadian amplitude to well capture circadian amplitude information with a strong genetic basis and clinical relevance, which can allow us to infer the causal relationship between circadian rhythms and cognitive functions.

Therefore, in this study we developed a new amplitude metric for circadian rhythms based on the accelerometer data and explored its clinical potential in two large-scale databases with different age groups. The workflow chart is shown in **Figure 1**. First, we proposed a pipeline to derive a new metric called **c**ircadian **a**ctivity **r**hythm **e**nergy (CARE) which can remove the influence of behavioral factors using spectral analysis approaches, and then we validated the new metric CARE with the melatonin amplitude. Secondly, we applied CARE to two accelerometer datasets with 1,703 adolescents and 92,202 adults, respectively, to examine the relationship between CARE and cognitive functions. Finally, we explored the causal relationship between CARE and cognitive performance through a genome-wide association study (GWAS) and MR causal analysis.

**Figure 1.**
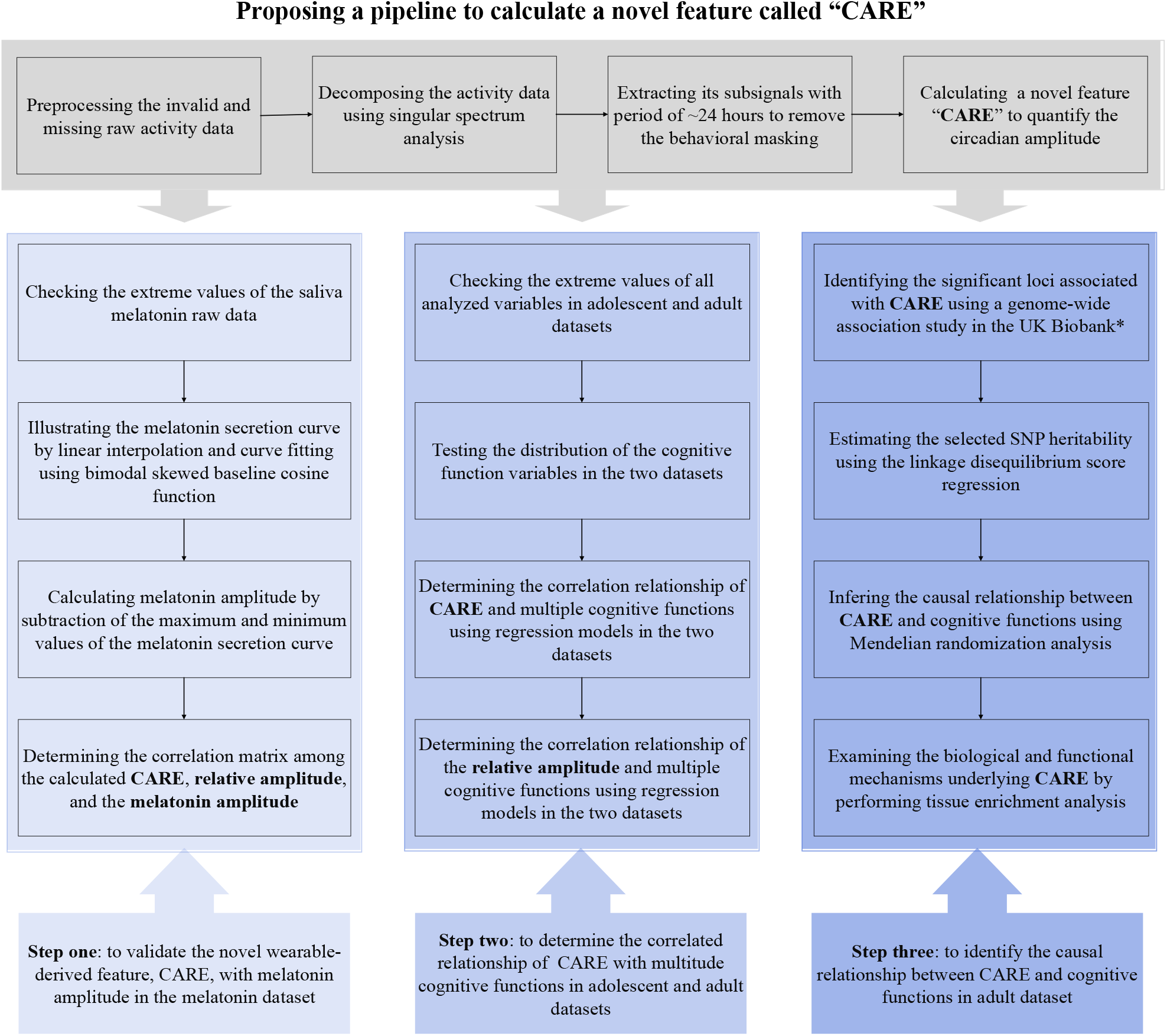
Workflow chart of the study. CARE: circadian activity rhythm energy *We also selected the significant loci that were associated with the relative amplitude. However, only three single nucleotide polymorphisms (SNPs) were found, the number of which was too small to conduct further causal analysis.

## Results

### A novel wearable-derived measure of circadian amplitude: CARE

In this paper, we developed a pipeline to extract a novel metric CARE (i.e., **c**ircadian **a**ctivity **r**hythm **e**nergy) to accurately quantify the circadian amplitude at a large scale with low costs. CARE can extract the circadian feature via acceleration signal decomposition, circadian sub-signal extraction, and energy estimation of the sub-signals from accelerometer data (details shown in **Figure 2**).

The accelerometer data can be considered as a composition of sub-signals with different frequencies. In order to characterize circadian rhythms with behavioral masking effects removed, we decomposed the data of activity counts and extracted the sub-signals with a period of ∼24 hours to represent the endogenous circadian oscillation. Among a list of signal decomposition approaches such as Fourier and wavelet analyses, we chose singular spectrum analysis (SSA, see a detailed description in **Figure 2c**) in our study for its data adaptive property[34, 35] and better performance compared to other methods such as Fourier and wavelet transform (detailed results can be found in **Supplementary Method**). Inspired by previous decomposition analysis on accelerometer data[23, 36], we used the relative energy of the circadian sub-signal with respect to the original signal as a novel estimate of the circadian amplitude. For a discrete-time signal x(t), energy is defined as the envelope of squared signal magnitude:

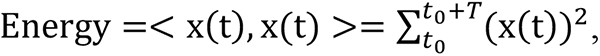

where *t*_0_ is the starting time point of x(t) and *T* is the time span of x(t). The unit of x(t) and energy are (activity counts)/minute and (activity count)^2^/minute^2^, respectively.

In summary, CARE is calculated using the following equation:

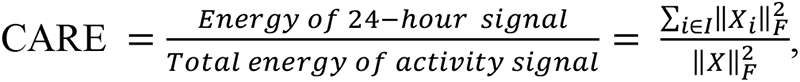

where 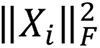 is Frobenius norm of the *i*th sub-signal, *I* is the subset of indices corresponding to the 24-hour signal, and 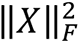 is Frobenius norm of the raw activity signal. It should be noted that CARE is expressed as a ratio ranging from 0 to 1 and is unitless. Furthermore, to effectively capture the dominant temporal scales and variability in the accelerometer data, and to obtain a reliable estimate of the autocovariance matrix feature, we required a minimum input data length of three days to calculate the CARE metric.

**Figure 2.**
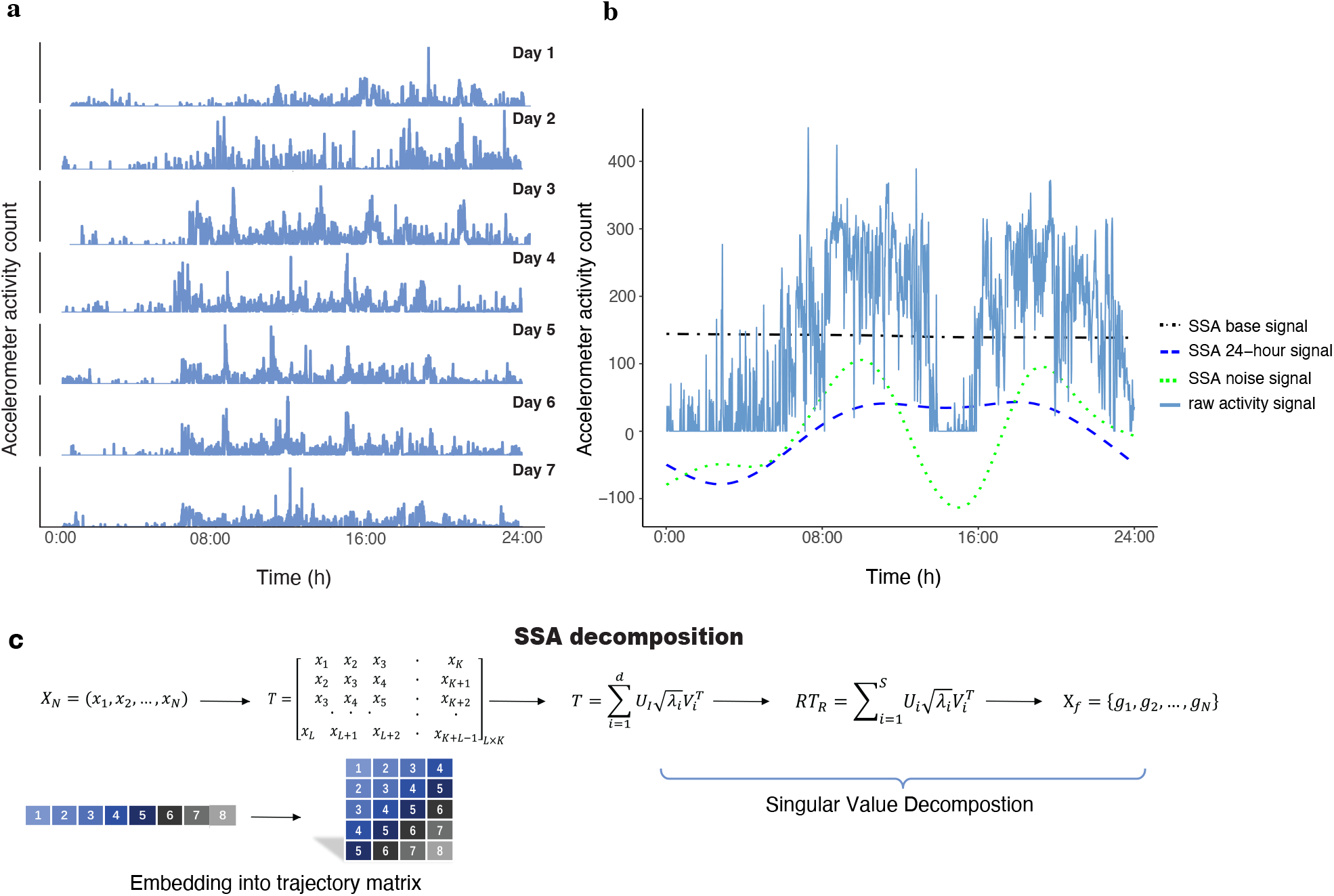
Pipeline to derive the novel measure circadian activity rhythm energy (CARE) from accelerometer data. (**a**) Actogram to show 7 days of wrist accelerometer activity of a single participant. (**b**) A visual representation of the raw activity signal and the singular spectrum analysis (SSA) decomposed signals, including the base signal (i.e., the first sub-signal indicating the non-periodic trend with the largest energy among all sub-signals), the 24-hour signal, and the behavioral noise signal (i.e., sum of the sub-signals with periods < 24 hours). We can obtain the raw activity signal by adding these three signals together. (**c**) The graphical display of SSA algorithm. Specifically, the activity time series X_!_ of length *N* could be decomposed using SSA as follows. First, we chose an appropriate window length *L* such that 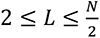. Then, X_*N*_ was transferred into a trajectory matrix with *K* lagged vectors of X_L_ as given by *T*, where *K* = *N – L +* 1. The trajectory matrix T was decomposed by singular value decomposition. By grouping the eigentriples and averaging the elements of reconstructed trajectory matrix along anti diagonals, we could get filtered time series represented by G_*N*_ = *g*_1_ *g*_2_ …, *g*_*N*_}.

**Table 1.**
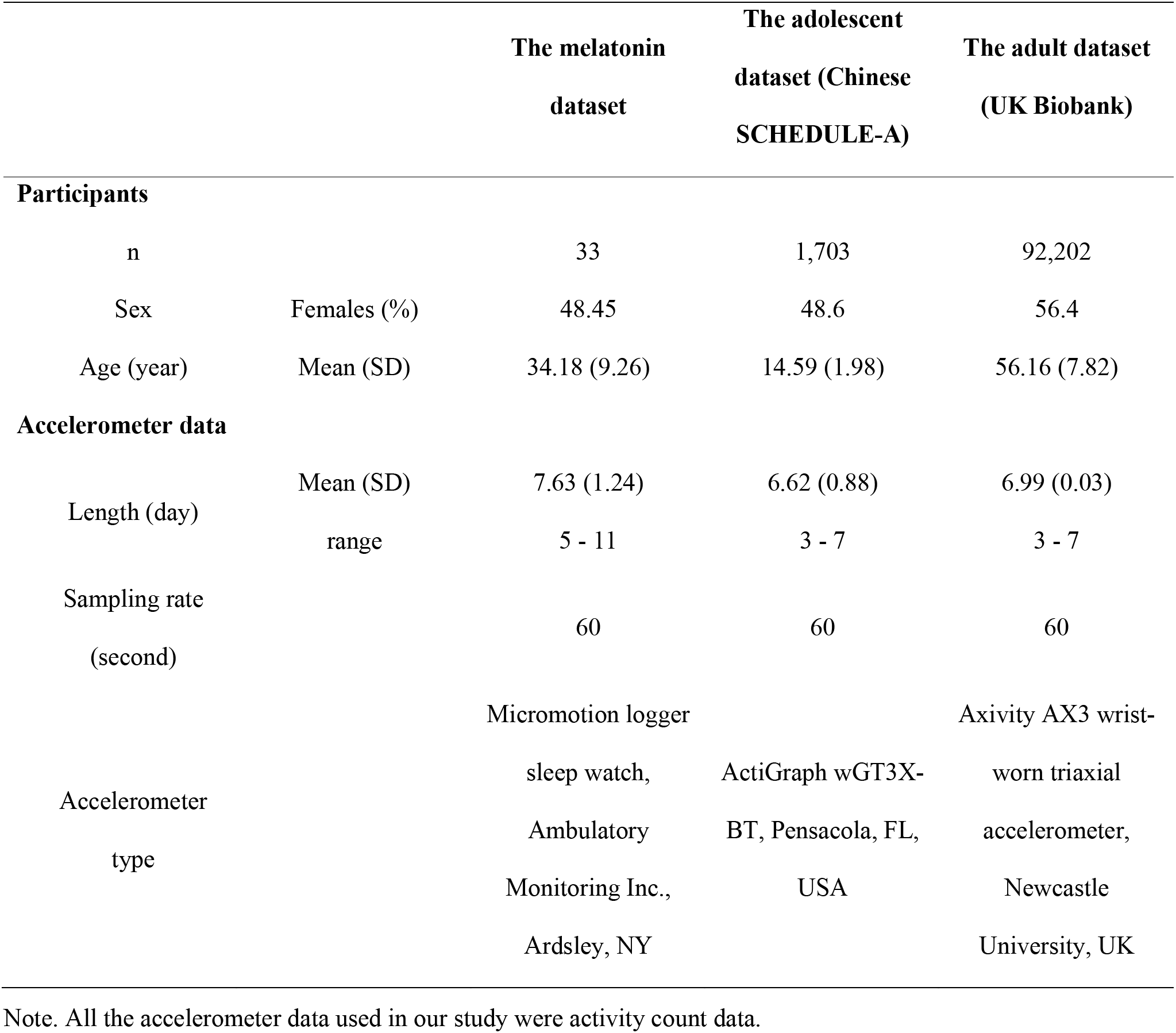
Description of the participants and the accelerometer data.

### Validate CARE with melatonin amplitude

We validated the novel feature CARE by examining its association with the melatonin amplitude in a dataset of 33 healthy participants aged 23 – 61 years, where accelerometer activity and melatonin were simultaneously collected (**Table 1**). Among the 33 participants, mean CARE was 0.26 (*SD* = 0.05; range 0.14 – 0.35), mean relative amplitude was 0.86 (*SD* = 0.07; range 0.67 – 0.96), and mean melatonin amplitude was 11.44 pg/ml (*SD =* 6.81; range 1.02 – 28.27). The correlation analysis revealed that the melatonin amplitude was only significantly associated with CARE (Pearson’s *r* = 0.46, *P =* 0.007; **Figure 3a**). On the other hand, we observed no significant association between melatonin amplitude and relative amplitude (Pearson’s *r* = 0.24, *P =* 0.19; **Figure 3**) or relative energy of behavioral noise signals (Pearson’s *r* = 0.07, *P =* 0.68; **Figure 3**). Moreover, our study found no significant association between age and sex with melatonin amplitude (*P* > 0.05; **Supplementary Table 1**). We also found that CARE accounted for 21.16% of the total variance of melatonin amplitude, whereas age and sex accounted for only 4.3%, and 0.03% of the variance, respectively (**Supplementary Table 1**).

**Figure 3.**
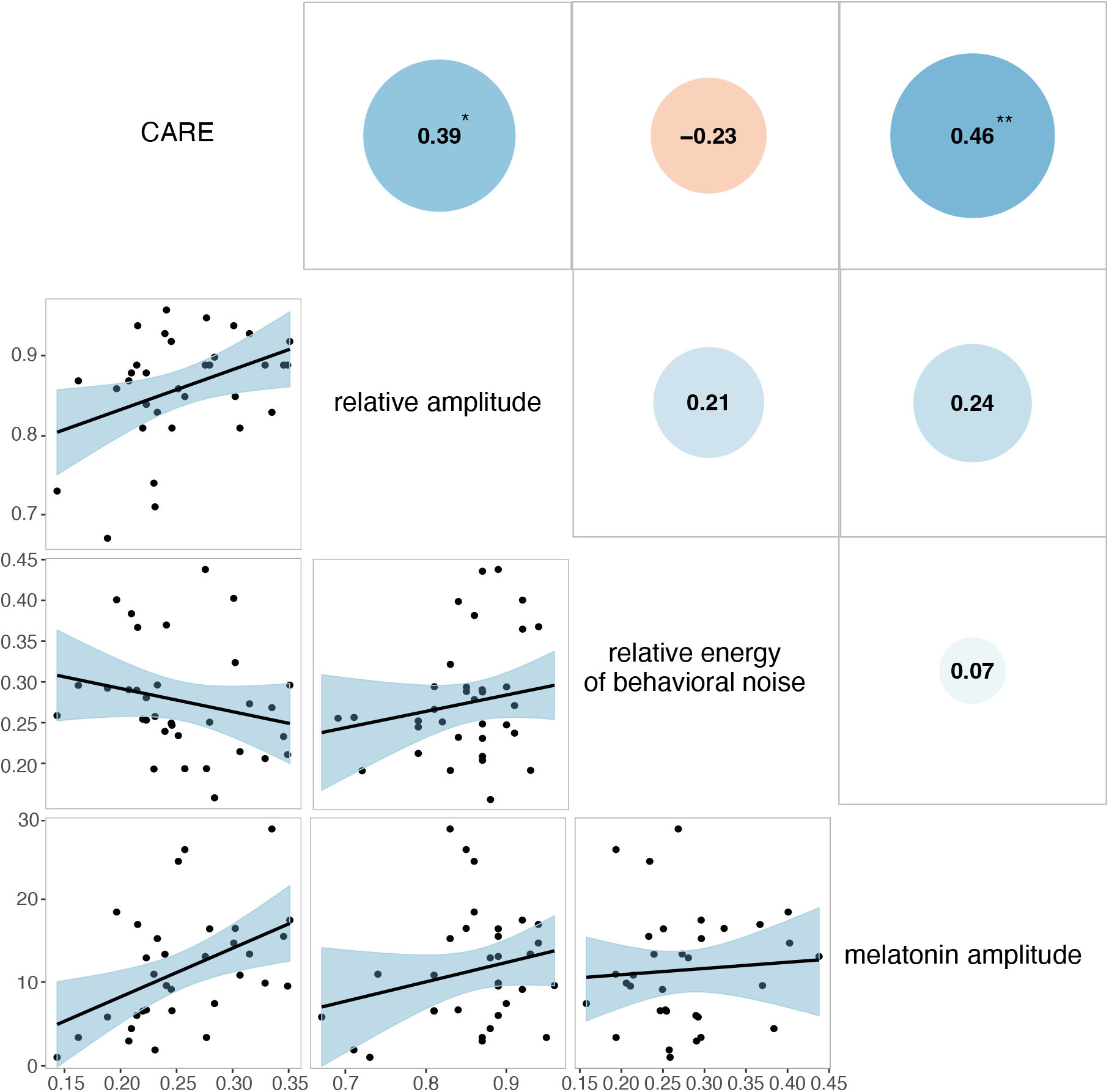
Correlations between wearable-derived CARE, relative amplitude, and relative energy of behavioral noise with melatonin amplitude. CARE: circadian activity rhythm energy. Scatter plots between CARE, relative amplitude, and relative energy of behavioral noise with melatonin amplitude in the melatonin study. The black lines indicate the linear regression fits, and the shaded areas represent the confidence intervals of the fitted mean values. The correlation coefficients annotated with an asterisk (*) indicate that the correlation is significant at *P* < 0.05 level, while two asterisks (**) indicate that the correlation is significant at *P* < 0.01 level.

### Correlation of CARE and cognitive outcomes in adolescents and adults

We applied CARE to two population-based accelerometer datasets with a total of 1,703 adolescents aged 10-19 years from China and 92,202 adults aged 39-70 years from UK. Characteristics of the participants and accelerometers data are presented in **Table 1**, and other analyzed variables are described in **Supplementary Tables 2 and 3**.

Mean CARE values were 0.10 (0.04) and 0.13 (0.04) for adolescents and adults, respectively. And mean relative amplitude values were 0.91 (0.05) and 0.87 (0.06) for adolescents and adults, respectively. Our results showed that CARE was significantly associated with relative amplitude in both populations (Pearson’s *r* = 0.41, *P* < 0.0001 for the adolescents; Pearson’s correlation *r*: 0.35, *P* < 0.0001 for the adults). Furthermore, we confirmed the internal consistency of CARE by analyzing the intra-subject versus inter-subject variance. The results showed that the intra-subject variability of CARE values was not significant in both adolescents (*P* = 0.18) and adults (*P* = 0.25), while the inter-subject variability was significant in both datasets (*P* < 0.0001, **Supplementary Table 4**). Moreover, we found significant between-group differences in CARE values among individuals with psychiatric disorders, such as bipolar affective disorder, schizophrenia, depression, and those in the control group (*P* = 0.02, **Supplementary Table 5**).

In the adolescent dataset, we used the Behavior Rating Inventory of Executive Function (BRIEF)[38] to assess adolescent cognitive functions in nature settings, which includes three summary indices, namely the Behavioral Regulation Index (BRI), the Metacognition Index (MI), and the Global Executive Composite (GEC). After adjusting confounding factors, the results showed that CARE had significant associations with GEC scores (*P* = 0.016), but it was not associated with BRI (*P* = 0.10) and MI scores (*P* = 0.03), (see detailed results in **Table 2**). However, no significant association was found between relative amplitude with any of the cognitive scores (*P* > 0.017) (see detailed results in **Supplementary Table 6**).

**Table 2.** Associations between CARE and cognitive functions in adolescents.

| Cognitive Scores | n | Coefficient | SE | *P* Value |
| --- | --- | --- | --- | --- |
| **BRI** | 1,703 | 19.86 | 11.89 | 0.10 |
| **MI** | 1,703 | 24.93 | 11.57 | 0.03 |
| **GEC** | 1,703 | 28.02 | 11.69 | 0.016 |

BRI = Behavioral Regulation Index, GEC = Global Executive Composite, MI = Metacognition Index, SE = standard error. Median regression models were used with adjusting age, sex, parental education level, family income, and primary caregiver. The significance level was set at $P < 0.017$ .

In the adult dataset, cognitive assessment was conducted via touchscreen cognitive tests, and four main tasks were considered, i.e., reaction time test assessing processing/reaction speed domain, fluid intelligence test assessing reasoning ability domain, pairs matching test assessing short-term memory domain, and prospective memory test assessing prospective memory. We found that CARE showed significant correlations with reasoning ability, short-term and prospective memory (*P* < 0.0001), but correlations were not significant for processing/reaction speed (*P =* 0.31). Details regarding the associations between CARE and cognitive scores are represented in **Table 3**. Though we found that relative amplitude was correlated with processing/reaction speed (*P <* 0.0001), it was not associated with reasoning ability, short-term and prospective memory (*P* > 0.013). Details regarding the associations between relative amplitude and cognitive scores are represented in **Supplementary Table 7**.

**Table 3.** Associations between CARE and cognitive functions in adults.

| Cognitive Scores | n | Coefficient / OR | SE | *P* Value |
| --- | --- | --- | --- | --- |
| **Processing/reaction speed** | 91,830 | -8.37 | 8.22 | 0.31 |
| **Reasoning ability** | 34,656 | 0.01 | 0.003 | <0.0001 |
| **Short-term memory** | 77,439 | 3.42 | 0.62 | <0.0001 |
| **Prospective memory** | 34,173 | 11.47 | 6.86 | <0.0001 |

OR = odds ratio, SE = standard error. Models were adjusted for age, sex, ethnicity, Townsend score, and the season when the participant started wearing the accelerometer. Linear regression for processing/reaction speed, ordinal logistic regression for reasoning ability and short-term memory scores, and logistic regression for prospective memory were employed. The significance level was set at $P < 0.013$ .

### Causal effects of CARE on cognitive functions

To determine the causal relationship between CARE and cognitive functions, we conducted a GWAS study of CARE followed with MR analysis in the adult sample, which contains filtered genetic data for 85,361 people.

#### 1) Genomic locus associated with CARE

One genomic locus on chromosomes 3 was found to correlate with CARE at the genome-wide significance level of 5 ×10^-8^, including 126 single nucleotide polymorphisms (SNPs) (**Figure 4**, lead variant in **Supplementary Table 8** and all significant associations in **Supplementary Data 1**). The quantile-quantile (QQ) plot checking for population stratification indicated proper control of population structure, with a slight deviation in test statistics compared to the null (**Figure 4**, λ_GC_ = 1.10). The heritability estimate for CARE was >11% (h^2^_SNP_ = 0.114, se = 0.007). This locus was reported to be associated with common executive functions[39], intelligence[40] and educational attainment[41] in the previous studies. However, only three SNPs were found to be associated with relative amplitude, which was too few for causal inference using MR methods (**Supplementary Figure 2** and **Supplementary Table 9**).

**Figure 4.**
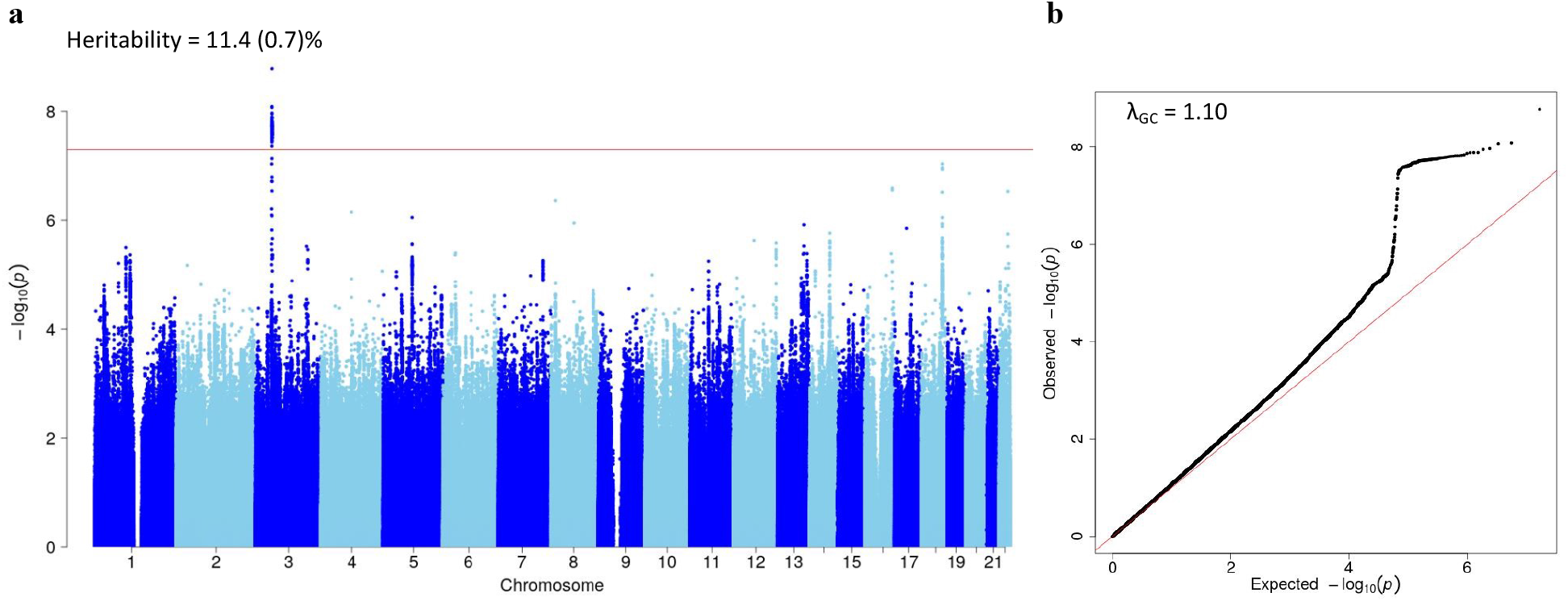
Manhattan and QQ plots for CARE in genome-wide association study in the adult sample (n = 85,361). (**a**) The Manhattan plot shows association test (-log_10_ *P* value on the y-axis against physical autosomal location on the x-axis). The red line represents the genome-wide significant locus (*P* < 5×10^−8^). Heritability estimates were calculated using LDSC tool. (**b**) The QQ plot identifies a slight inflation (λ_GC_ = 1.10) in the test statistic.

#### 2) Causal relationship between CARE and cognitive phenotypes

To investigate whether CARE has a causal effect on cognitive outcomes, we performed MR analyses on significant correlation pairs, i.e., CARE with reasoning ability, short-term and prospective memory (**Table 3**). Of the 126 CARE-related SNPs, a total of 109 variants were used as instrumental variables to conduct the MR analyses. Using the weighted median method, we found that the genetically predicted CARE had a significant causal effect on reasoning ability (*β* = −59.91, *P* < 0.0001), short-term (*β* = 7.94, *P* < 0.0001) and prospective memory (*β* = 16.85, *P* < 0.0001). Sensitivity analyses including inverse variance weighted (IVW)[42], MR-Lasso[43], the mode-based estimate (MBE)[44], and constrained maximum likelihood-based method (MR-cML)[45] all showed consistent results with weighted median methods (**Supplementary Table 10**).

#### 3) Single-tissue and cross-tissue transcriptome-wide association analysis

Single-tissue enrichment analysis identified 120 unique genes associated with CARE (*P <* 2.9×10^-6^) in 44 GTEx tissues (**Supplementary Data 2**). Among them, APEH was associated with CARE in 18 tissues and was found to correlate with blood protein measurements in previous GWAS studies[46]. TRAIP was detected in 11 tissues and was reported to be associated with intelligence[39], mental disorders[47, 48], insomnia[49], blood protein measurement[50, 51], and body mass index[52]. MST1R was associated with CARE in 9 tissues and is related to physical activity measurements[53], intelligence[54], and body mass index[55]. These correlation results also emphasized shared genetic architecture between the central nervous system, the metabolic system, and the circadian system. Furthermore, a total of 15 significant genes were detected using joint-tissue tests for gene-CARE associations across tissues (*P <* 2.9×10^-6^; **Supplementary Table 11**).

## Discussion

In the present study, we proposed a pipeline to derive a novel wearable-based feature (i.e., CARE) to characterize circadian amplitude and applied it to identify the association between circadian amplitude and cognitive functions in two large-scale datasets with different age groups. We found a significant association between CARE and melatonin amplitude (a reliable measure of circadian amplitude), while relative amplitude (a commonly used amplitude of the activity) was not. Findings that CARE was associated with a multitude of cognitive outcomes in adolescents and adults demonstrated that CARE could be a clinically meaningful circadian feature derived from objective accelerometer records. Furthermore, we identified one genetic locus with 126 SNPs associated with CARE, and provided the first direct evidence of the causal relations between CARE with reasoning and memory abilities in an adult sample.

Our study found a moderate correlation between the new feature CARE and melatonin amplitude in general population under natural settings. The use of CARE can effectively eliminate the influence of behavioral noise on the assessment of circadian amplitude using the accelerometer data, as supported by the lack of significant association between behavioral noise signals and melatonin amplitude. Notably, we calculated CARE values using accelerometer activity data of at least 3 days, making it a summary statistic across multiple days for each individual. This computation approach enhances the stability of CARE values and reduces the intra-subject variability. Nonetheless, it is important to note that the CARE values may be impacted by data obtained from various accelerometer devices, which could potentially affect their comparability. Furthermore, the range of CARE values observed in the melatonin dataset is relatively narrow, which is likely due to the limited range of observed melatonin amplitude, as the maximum melatonin amplitude can be up to 84 pg/ml in healthy adults[56]. Therefore, it would be beneficial to investigate whether a broader range of melatonin amplitude leads to a wider range of CARE values in the future research.

The findings that age and sex were not significantly associated with melatonin amplitude may contradict with previous research that melatonin levels typically decrease with age starting from middle age and that women tend to have higher melatonin amplitude than men [57–59]. This might be due to the limited number of middle-aged and elderly participants (age > 40) and the limited sample size in our dataset. Future research shall recruit larger samples, especially from the elderly adults, to further examine age and sex differences.

Our study demonstrated that CARE and relative amplitude may assess different aspects of circadian rhythmicity. Although they are all derived from accelerometer data, CARE was designed to reflect the strength of the core circadian clock located in the SCN, and relative amplitude is more of a metric to quantify the highest/lowest disruption of rest-activity rhythms. The identification of a sufficient number of CARE-associated variants can enable us to perform causal inference through MR analysis, which is not possible with relative amplitude. In addition, the SNP heritability of CARE accounted for a higher proportion of population variance than relative amplitude in the adult sample (UK Biobank cohort). These results further supported CARE as a clinically meaningful feature. Future research is still warranted to systematically compare and assess the relative contribution and relevance of CARE and relative amplitude, particularly in the context of health and disease.

Our study observed a differential association between CARE and cognitive functions in adolescent and adult groups. In adults, we observed a correlation between CARE and the problem-solving component of cognitive functions, specifically reasoning and memory domains. However, in adolescents, this association was not present. This discrepancy may indicate the existence of a compensatory mechanism in adolescents that counterbalances the impact of circadian rhythm impairments on problem-solving ability. More research is needed to confirm our hypothesis in the future.

In the present study, we demonstrated a causal effect of CARE on reasoning and memory abilities in adults, and we also provided evidence for the shared genetic architecture of circadian rhythms with neurological function[9]. The genome-wide significant locus was found to be associated with common executive functions[39], intelligence[40], and educational attainment[41]. Furthermore, results from the single-tissue and cross-tissue transcriptome-wide association analyses further confirmed the CARE’s strong biological basis and highlighted the important role of circadian rhythms on the central nervous system, which is in line with previous findings from the other UK Biobank studies[60].

In conclusion, the new feature CARE that we derived from accelerometer data, is closely related to the melatonin amplitude in natural settings, and also serves as a meaningful metric for a wide range of cognitive functions in general adolescents and adults. Future studies with large sample size and different protocols such as forced desynchrony settings and shift work should be conducted to validate the new feature CARE and to confirm its causality on other health-related outcomes.

## Methods

### The melatonin dataset

#### Participants

This dataset recruited 33 healthy adults (17 male and 16 female) aged 23-61 years in January 2023. Prior to enrollment, all participants underwent screening to exclude the use of sleep aid supplements (e.g., melatonin), a history of shift work, and sleep disorders. The participants were asked to complete a range of demographic, lifestyle, and health related questionnaires, to wear an accelerometer, and to collect saliva samples with 5 times. This protocol was approved by the Shanghai Children’s Medical Center’s Human Ethics Committee (SCMCIRB-K2021070-2) and Shanghai Jiao Tong University School of Medicine (SJTUPN-202301). All participants provided digital written informed consent.

#### Accelerometer data collection and processing

The participants were asked to wear an accelerometer (Micromotion logger sleep watch, Ambulatory Monitoring Inc., Ardsley, NY) on their non-dominant wrist for nine consecutive days, meanwhile, they were required to keep a diary recording bedtime, rise time, and off-wrist period on each day. They were informed not to remove the device unless engaged in special activities (e.g., showering), and to re-wear it immediately after the activities were finished. We defined the daytime period as 08:00-20:00 and the nighttime period as 20:00-08:00. According to the diary recording for the off-wrist periods, we defined a criterion that at least 15 min of consecutive zero counts were identified nonwear periods in period 08:00-23:00, and at least 3 hours of consecutive zero counts were identified nonwear periods in the remaining period. We also considered that the valid data on each day should meet at least 20 hours (i.e., more than 83% of the whole 24 hours) of wearing time without more than three consecutive hours of missing data. If less than 3 hours of missing data existed, we used the mean of the corresponding time points on the other days to fill the gaps. After checking, if there were still missing values (often due to the missing data being at a fixed time every day, so it could not be filled using the mean value), the average of physical activity counts around the missing area (i.e., about 6 epoch) was used to fill. After preprocessing the invalid and missing raw data, each individual included should have at least three valid days of data, and the activity counts with 1-min resolution were used to calculate the new feature CARE using the above-proposed novel pipeline and to compute the commonly used feature relative amplitude by the difference between the average activity level of an individual during the most active consecutive ten hours (M10) and the least active consecutive five hours of the day (L5).

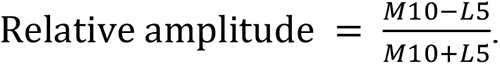

#### Saliva melatonin data collection and processing

We used the Salivettes^®^ (Sarstedt, Nümbrecht, Germany) to collect saliva. On the eighth and ninth day of wearing the accelerometer, the participants were asked to collect 2 ml saliva samples 5 times (i.e., 9 am, 15 pm, 21 pm, 3 am, and 9 am of the next day; time window ±1 hour) across 24 hours, of which the saliva sample at 21 pm and 3 am were required to be collected under the dark light (light intensity < 30 lux) if the participants usually went to sleep at that time. The participants were informed to follow their usual sleep schedule and don’t intake caffeine, alcohol, and fatty foods on the sampling day, keep their mouth clean (e.g., food intake restriction) in the 30 min prior to sampling, and do not swallow saliva while chewing the swab. They were also informed to store each sample immediately in the freezer layer of the refrigerator after collection, and to transfer all of the samples to the laboratory using ice packs. Subsequently, after centrifugation at 4℃ for 10 min at 3000 rpm, the supernatant saliva samples were collected into 2 ml EP tube and stored at -80℃ until use.

Saliva melatonin was measured by liquid chromatography-tandem mass spectrometry, which can quantify the range of the melatonin from 1 to 1000 pg/ml, and only use small volume of the saliva samples (100 µl). Detailed measurement was similar to the quantification of melatonin in human milk[61]. For more information, please contact the corresponding author. After quantification of saliva melatonin levels, we checked the extreme values, and we excluded the values less than 2 pg/ml which collected at 3 am (time window ±1 hour). And then melatonin profiles were obtained by linear interpolation and curve fitting using skewed baseline cosine function (SBCF) (**Supplementary Figure 3**), a robust model for melatonin estimation when there were limited time points in the collection of melatonin[62].

### The adolescent dataset

#### Participants

The Shanghai Children’s Health, Education, and Lifestyle Evaluation-Adolescent (SCHEDULE-A) is a population-based cross-sectional study designed to investigate the physical and mental health of adolescents (10-19 years). A total of 1,703 adolescents from a subsample of SCHEDULE-A (Shanghai city, China) were utilized for this study. Two data acquisition schemes, an online questionnaire survey, and offline accelerometer wearing, were performed simultaneously to collect adolescents’ socio-demographic information, lifestyle behaviors, physical and mental health outcomes. Details of the protocol can be found in one prior published study[63]. This study was approved by the Human Ethics Committee of the Shanghai Children’s Medical Center according to the Declaration of Helsinki (SCMCIRB-K2018103). Written informed consent was obtained from all participants.

#### Accelerometer data collection and processing

Participants were asked to wear an accelerometer (ActiGraph wGT3X-BT, Pensacola, FL, USA) on the non-dominant wrist for seven consecutive days. The other requirements including device wearing and diary recording were similar to the melatonin dataset. Using the same way to preprocess the raw data, and then CARE and relative amplitude were calculated.

#### Measurement of cognitive functions

The Chinese version of the BRIEF was used to assess adolescent cognitive functions in nature settings[64]. The BRIEF has 86 items, each one is rated on a three-point Likert-type scale, including never, sometimes and often, and parents of the participants were asked to select the most suitable answer for their adolescents in the past 6 months. After cleaning the raw data[63], three indexes were calculated: 1) the BRI, represents the ability to modulate behavioral and emotional responses appropriately; 2) the MI, evaluates the ability to solve problems, including initiating activities and tasks, holding information in mind for purpose of completing a task, anticipating future events, monitoring the effects of one’s behaviors on others and keeping track of daily assignments; 3) the GEC, is a composite measure of all the cognitive functions mentioned above. Values of BRIEF scores reflect the extent of impairment of the corresponding cognitive functions, and higher scores indicate greater impairments.

#### Covariates

One of the participants’ parents was asked to complete a questionnaire about their social-demographic characteristics, i.e., adolescents’ age, sex, primary caregiver (parents/grandparents or others), family income (< 50,000 CNY/year, 50,000 to 150,000 CNY/year, ≥ 150,000 CNY/year), and parental education level (below high school, high school, college and above).

### The adult dataset

#### Participants

The UK Biobank cohort is a general longitudinal study with over 500,000 participants recruited from the United Kingdom (UK). In the current study, we selected 92,202 participants who provided qualified accelerometer data for at least three days after data preprocessing and who had complete records in age and sex. The demographic, lifestyle, health, mood, cognitive, and physical conditions of the participants were collected at 22 assessment centers. The UK Biobank has received ethical approval from the North-West Research Ethics Committee (11/NW/03820) and all participants gave written informed consent.

#### Accelerometer data collection and processing

Participants were asked to wear an accelerometer (Activity AX3 wrist-worn triaxial accelerometer, Newcastle University, UK) for seven consecutive days. The accelerometer raw data were preprocessed similarly to the above melatonin and adolescent datasets. Participants were also excluded if their data were identified by UK Biobank as poorly calibrated (n = 11) or unreliable (unexpectedly small or large size, n *=* 4,692). Participants who provided accelerometer data for less than 72 hours or who did not provide data for all one-hour periods within the 24-h cycle (n *=* 4,465) were also excluded from further analyses. Similarly, after cleaning the raw data, CARE and relative amplitude were calculated.

#### Measurement of cognitive functions

Four tests were used to assess adult processing/reaction speed, reasoning ability, short-term memory, and prospective memory. 1) The processing/reaction speed was assessed by the reaction time test. During the test, participants were shown 12 pairs of images of cards for 12 rounds, and they pressed the button on the touchscreen when the two cards are the same. The processing/reaction speed was then calculated by the mean reaction time of the correct matches over the last 7 rounds after the exclusion of times below 50ms and above 2,000ms. Due to negative skewness in reaction time, we performed the analyses using both the raw and log-transformed data form. Since the results were equivalent, only the results of raw reaction time were reported for ease of interpretation. 2) Reasoning ability was measured by the fluid intelligence tests. Participants were asked to answer 13 fluid intelligence questions within 2 minutes and the total number of correct answers to these questions was recorded as the fluid intelligence score. 3) Short-term memory was assessed by the pairs matching tests. Participants were asked to memorize as many pairs of cards as possible within the time the cards were shown. Then the cards were turned face down and the participants were asked to match the cards. The test was conducted in two rounds, with 3 and 6 pairs of cards presented in each round. The outcome measure used in this study was the number of incorrect matches in rounds, as suggested by the UK Biobank website. 4) Prospective memory was assessed by the prospective memory test. Before the other touchscreen cognitive tests, the participant was shown the message “At the end of the games we will show you four colored shapes and ask you to touch the Blue Square. However, to test your memory, we want you to actually touch the Orange Circle instead”. The results were divided into 3 categories, 0 for instruction not recalled, 1 for correct recall on the first attempt, and 2 for correct recall on the second attempt, respectively. The outcome measure used in this study was the number of attempts before getting the correct recall.

#### Covariates

Participants provided data on demographic characteristics including age, sex, ethnicity, educational attainment, and lifestyle at the baseline assessment. Townsend’s social deprivation score is defined based on the postcode of residence, with negative scores reflecting relatively greater affluence. The season when the participant started to wear the accelerometer was derived from raw accelerometer records, i.e., spring for March to May, summer for June to August, autumn for September to November, and winter for December to February according to the UK Meteorological Office definitions.

### Statistical analysis

The characteristics of the three datasets were described by mean (*SD* and range), and frequency (%) for continuous and categorical variables, respectively. To address our purpose, the following three steps were conducted after calculating the new feature CARE according to the above-proposed pipeline (**Figure 1**).

***Step one***: to validate the novel wearable-derived feature CARE

We firstly calculated the new feature CARE using the above-proposed pipeline from accelerometer data. Secondly, using the same pipeline, we also derived the relative energy of behavioral noise signals (sub-signals with a period less than 24 hours) from accelerometer activity data:

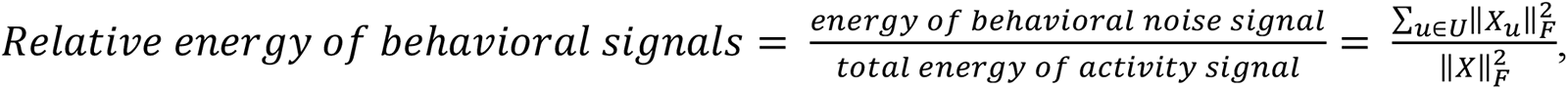

where 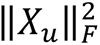 is Frobenius norm of the *u*th sub-signal, *U* is the subset of indices corresponding to the behavioral noise signal (sub-signals with a period < 24h), and 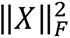 is Frobenius norm of the raw activity signal. Thirdly, we obtained the melatonin profiles by linear interpolation and curve fitting using skewed baseline cosine function (SBCF), a robust model for melatonin estimation when there were limited time points[62]; Fourthly, the melatonin amplitude was measured by subtraction of the maximum and minimum values of the melatonin secretion curve; and finally, we conducted Pearson correlation analyses between CARE, relative amplitude, and the relative energy of behavioral high-frequency signals with melatonin amplitude. Additionally, we examined the associations between CARE and melatonin amplitude while adjusting for age and sex.

***Step two***: to determine the correlation relationship of CARE with multitude of cognitive functions in adolescents and adults

Prior to performing the association analysis, we evaluated the internal stability of CARE values through two distinct analyses. Firstly, we randomly selected a subset of 1,000 individuals each from the adolescent and adult datasets who had at least six days of accelerometer data. We then calculated the CARE values using the first three days of recordings (repetition 1) and the fourth to sixth days of recordings (repetition 2), respectively. An Analysis of Variance (ANOVA) was then performed to compare the intra-subject and inter-subject variability of CARE values. Secondly, we conducted another ANOVA to explore the within-group and between group variability of CARE values in bipolar affective disorder (n = 147), schizophrenia (n = 42), depression (n = 2,252), and control groups (n = 36,315) in the adult dataset. Specifically, the control group comprised individuals who did not currently have the above-mentioned psychiatric diseases, did not have a history of mental disease, did not take psychiatric medication, did not receive any professional help for mental distress, and had not suffered from mental distress.

We then performed different regression models to determine the relationship of CARE with multitude of cognitive functions in two datasets. Specifically, in adolescents, because all the three BRIEF indices (i.e., BRI, MI, and GEC) were skewed distributed, median regression analysis was used with adjustment for the demographic variables (i.e., age, sex, parental education level, family income, and primary caregiver); and in adults, linear regression model was performed to assess the associations between CARE and processing/reaction speed, ordinal logistic regression model was employed to investigate the associations between CARE and count outcome variables (reasoning ability, and short-term memory), while logistic regression model was conducted to test the correlation between CARE and the binary cognitive measures of prospective memory. Similarly, models were adjusted for age, sex, ethnicity, Townsend score, and the season when the participant started wearing the accelerometer. To maximize sample sizes, available-case analyses were employed, and sample sizes for each model were reported. Moreover, the analyses mentioned above were also performed on relative amplitude for the purpose of comparison.

All analyses were performed in R 4.1.2, and the threshold for significance was set at *P* < 0.05/3 = 0.017 (3 cognitive outcomes) and *P* < 0.05/4 = 0.013 (4 cognitive outcomes) to correct for multiple testing according to Bonferroni’s approach in adolescent and adult dataset analyses, respectively.

***Step three***: to identify the causal relationship between CARE and cognitive functions in the adult dataset.

#### 1) Identification of loci associated with CARE

Before the genetic analysis, we excluded SNPs with Minor Allele Frequency < 0.1%, Hardy-Weinberg equilibrium < 1 × 10^0>%^, and imputation quality score (UKBB information score) < 0.8. Furthermore, for related individuals (first cousins or closer), one individual from each pair of related individuals was randomly selected for inclusion in the analysis, and non-white individuals were also removed from the analysis. After data filtering, a total of 85,361 samples remained for further analysis. To identify genetic variants associated with CARE, we performed a GWAS analysis using PLINK[65]. Linear regression models were adjusted by age, sex, and the first 20 genetic principal components, and the genome-wide significance level was set at 5 × 10^−8^. To estimate SNP heritability (h^2^_SNP_), the Linkage Disequilibrium Score Regression (LDSR) was applied to the GWAS summary statistics of CARE with the LDSC tool[66, 67]. Furthermore, we identified one significant locus that was associated with the relative amplitude, however, only three SNPs were found, which was just too little that we couldn’t conduct the further causal analysis.

#### 2) Causal relationship between CARE and cognitive functions

To investigate the causal relationship between CARE and cognitive functions, we conducted MR analyses using the R package “MendelianRandomization”[68]. In order to exclude instrumental variables with weak power, we pruned CARE-related SNPs and finally determined 109 SNPS as instruments. To avoid inflation of associations caused by the sample overlap, genetic effect sizes for reasoning ability, short-term and prospective memory were obtained from GWAS analysis after excluding all individuals used in the GWAS analysis of CARE (n = 360,885). In the MR analyses, the weighted median approach[69] was used as the main analysis due to its robustness to pleiotropy [70]. Furthermore, we applied inverse variance weighted (IVW)[42], MR-Lasso[43], the mode-based estimate (MBE)[44], and constrained maximum likelihood-based method (MR-cML)[45] to perform sensitivity analyses. The beta coefficients were reported and Bonferroni-adjusted *P* values less than 0.017 (3 outcomes) were considered evidence of associations.

#### 3) Single-tissue and Cross-tissue Transcriptome-wide Association Analysis

In addition, we examined the biological and functional mechanisms underlying CARE by performing transcriptome-wide analysis in 44 tissue types in GTEx v7. We identified single-tissue and cross-tissue gene-CARE associations based on GWAS summary statistics and expression quantitative trait loci (eQTL) information, using UTMOST[71]. The Bonferroni-corrected threshold for significance in UTMOST is 2.9 × 10^−6^ for 17,290 genes.

## Data Availability

The melatonin and adolescent datasets used in the current study are not publicly available but are available from the corresponding author (Fan Jiang) on reasonable request. The UK Biobank data that support the findings of this study are available from the UK Biobank project but restrictions apply to the availability of these data, which were used under license for the current study (application number: 57947), and so are not publicly available.

## Code Availability

Codes for calculating CARE were publicly available at https://github.com/ShuyaCui/CARE.

## Acknowledgements

The study was supported by the National Key R&D Program of China (2018YFC0910500), National Natural Science Foundation of China (82073568), Shanghai Jiao Tong University (YG2016ZD04), Shanghai Municipal Health Commission (GWIV-36 and GWV-10.1-XK07), SJTU-Yale Collaborative Research Seed Fund, the Neil Shen’s SJTU Medical Research, and Innovative Research Team of High-level Local Universities in Shanghai (SHSMU-ZDCX20211900). The melatonin and adolescent datasets were kindly provided by the Shanghai Children’s Medical Center. The authors are grateful to all members for contributing data to the two databases. This research was also conducted with the UK Biobank resource, under application 57947. We thank all individuals involving in the UK Biobank study. Additionally, we extend our gratitude to Du Jingyu for his assistance in refining the language of the manuscript.

## Author contributions

Conceptualization: CS, LQ; Data curation: CS, LQ; Formal analysis: CS, GY; Funding acquisition: LH, JF; Investigation: CS, LQ, ZY; Methodology: CS, LQ, ZH, LX; Project administration: ZY, JF; Resources: LH, JF; Supervision: LH, ZH, WX, LX, JF; Validation: CS, LQ, GY; Visualization: CS; Writing——original draft: CS; Writing——review & editing: CS, LQ, GY, ZY, LH, ZH, WX, LX, JF. All authors read and approved the final manuscript.

