## Supplementary method, figures and tables for "CARE: a novel wearable-derived feature linking circadian amplitude to human cognitive functions"

**Determining a suitable method of signal decomposition.**

We have used data from the melatonin dataset to compare the correlations between melatonin amplitude and CARE values derived from three signal decomposition methods: the Fourier transform (FFT), discrete wavelet transform (DWT), and singular spectral analysis (SSA). We found that SSA-derived CARE value was significantly associated with melatonin amplitude (Pearson's $r$ = 0.48, $P$ = 0.005), but not FFT-derived (Pearson's $r$ = 0.20, $P$ = 0.26) and DWT-derived CARE values (Pearson's $r$ = 0.15, $P$ = 0.41). Besides, unlike FFT and DWT, the SSA does not require a fixed base function for signal decomposition, which makes it entirely data-driven and flexible to use in practice. Thus, we chose SSA as the decomposition method in our pipeline.

### Supplementary Figures

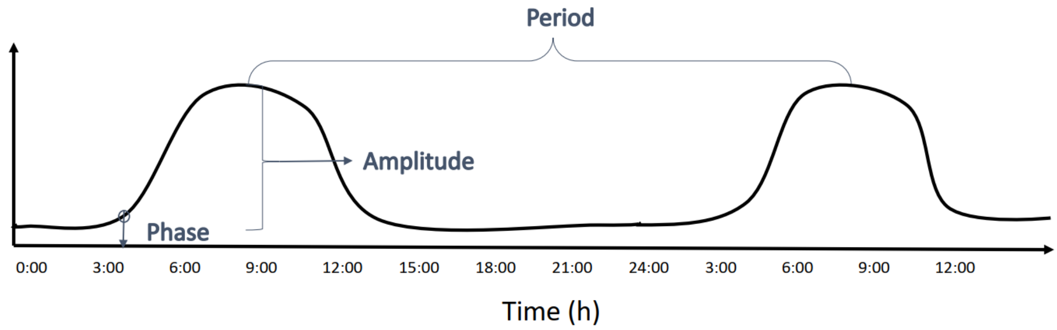


**Supplementary Figure 1. Illustration of period, phase and amplitude of circadian rhythms.**

Period is the length of a cycle, specifically, it is the time interval between two reference points within a recurring wave (for instance, between hormonal peaks). Phase is defined as the timing of a reference point in the cycle relative to a fixed event. In relation to the melatonin secretion cycle, for example, dim light melatonin onset (DLMO) is the gold standard for the phase of melatonin rhythms. Amplitude is defined as difference between crest and trough values of the cycle. In relation to the hormonal cycle, it would be the difference between the trough and peak hormone levels within a time period (i.e., 24 hours).

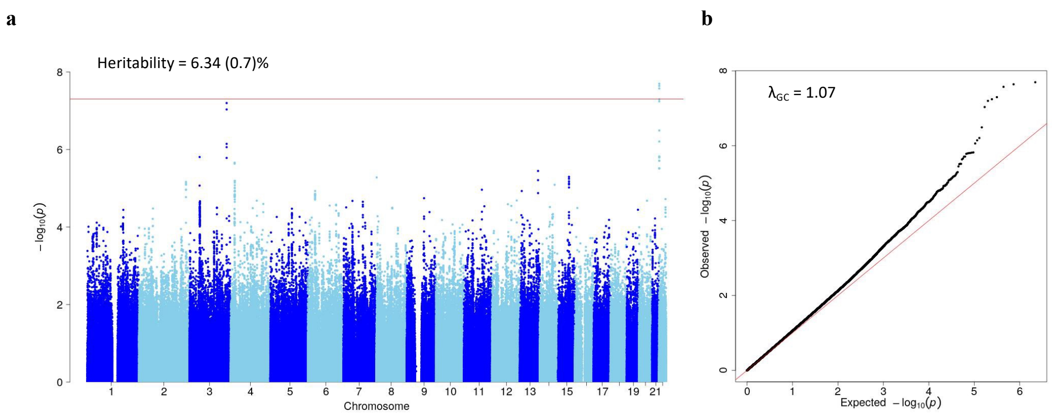


**Supplementary Figure 2. Manhattan and QQ plots for relative amplitude associated-SNPs in the adult dataset (UK Biobank) (n = 85,361).**

(**a**) The Manhattan plot shows association test (-$\log_{10}$ *P*-value on the y-axis against physical autosomal location on the x-axis). The red line represents genome-wide significance ($P < 5\times10^{-8}$). Heritability estimate was calculated using LDSC tool. (**b**) The QQ plot identifies a slight inflation ($\lambda_{GC} = 1.07$) in the test statistic.

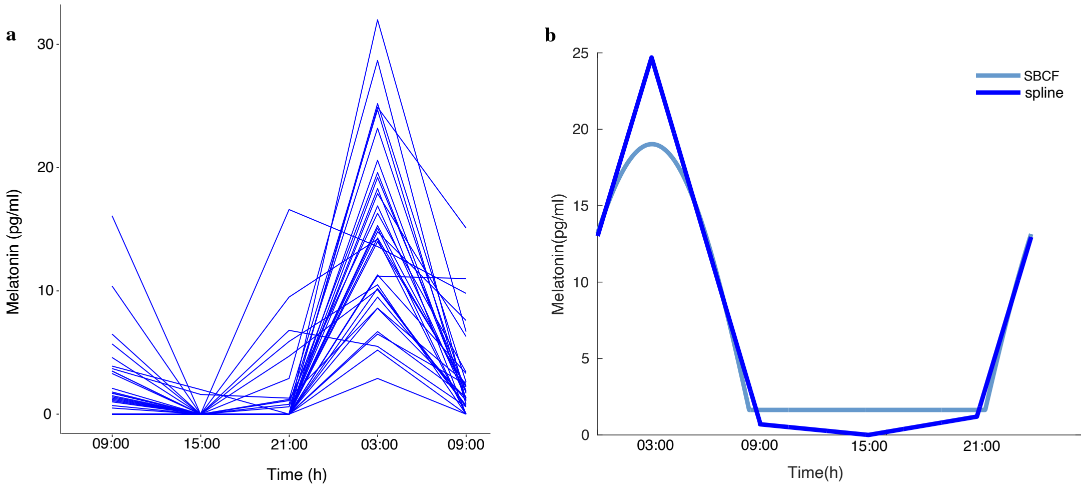


**Supplementary Figure 3. The melatonin profiles in the melatonin dataset.**

(**a**) Subject-level melatonin profiles (observed values) in the melatonin dataset. Saliva melatonin were collected for five times (i.e., 9 am, 15 pm, 21 pm, 3 am, and 9 am of the next day; time window ± 1 hour) during 24 hours for each participant. (**b**) Illustration of the daily melatonin profile of an individual. Five melatonin sample points were first linear interpolated and then fitted with the skewed baseline cosine function (SBCF) model to get the melatonin secretion curve. Melatonin amplitude was calculated by subtraction of the maximum and minimum values of the melatonin profiles.

### Supplementary Tables

#### Supplementary Table 1. Associations between CARE and melatonin amplitude in the melatonin dataset.

| Covariates | Coefficient | SE | *P* value | Variance explained |
|---|---|---|---|---|
| CARE | 58.38 | 20.24 | 0.007 | 21.16% |
| age | -0.15 | 0.12 | 0.21 | 4.3% |
| sex | -0.22 | 2.15 | 0.92 | 0.03% |

CARE = circadian activity rhythm energy, SE = standard error. Linear regression analysis was performed to examine the association between CARE and melatonin amplitude, adjusted for age and sex in the melatonin dataset. The significant level was set as $P < 0.05$.

**Supplementary Table 2. Description of the analyzed variables in the adolescent dataset (Chinese SCHEDULE-A).**

| Variables | Mean ± SD / n(%) |
|---|---|
| **Device-derived features of circadian amplitude** | |
| CARE | 0.10 ± 0.04 |
| Relative amplitude | 0.91 ± 0.05 |
| **Cognitive scores** | |
| BRI | 48.86 ± 9.22 |
| MI | 51.60 ± 9.80 |
| GEC | 50.63 ± 9.71 |
| **Other confounders** | |
| Parental education level | |
| *Lower than high school* | 426 (25.0) |
| *High school* | 482 (28.3) |
| *College or higher* | 795 (46.7) |
| Income level | |
| *<50,000 RMB* | 432 (25.4) |
| *50,000 ~ 150,000 RMB* | 765 (44.9) |
| *⩾150,000 RMB* | 506 (29.7) |
| Main caregiver | |
| *parents* | 1562 (91.7) |
| *grandparents or others* | 141 (8.3) |

BRI = Behavioral Regulation Index, CARE = circadian activity rhythm energy, GEC = Global Executive Composite, MI = Metacognition Index, SD = standard deviation.

**Supplementary Table 3. Description of the analyzed variables in the adult dataset (UK Biobank).**

| Variables | Mean ± SD / n(%) |
|---|---|
| **Device-derived features of circadian amplitude** | |
| CARE | 0.13 ± 0.04 |
| Relative amplitude | 0.87 ± 0.06 |
| **Cognitive scores** | |
| Processing/reaction speed | 545.50 ± 105.36 |
| Fluid intelligence scores | 7.58 ± 2.07 |
| Reasoning ability | 4.89 ± 0.72 |
| Prospective memory results (number of attempts) | |
| *1* | 29455 (86.2) |
| *2* | 4735 (13.9) |
| **Other confounders** | |
| Townsend scores | -1.73 ± 2.82 |
| BMI (kg/m$^2$) | 26.71 ± 4.53 |
| Average daily activity intensity | 27.99 ± 8.24 |
| Ethnicity | |
| *White* | 89045 (96.6) |
| *Non-white* | 3157 (3.4) |
| Qualification | |
| *Below college* | 39587 (42.9) |
| *College or higher* | 52070 (56.5) |
| Smoking status | |
| *Never* | 52532 (57.0) |
| *Previous* | 33090 (35.9) |
| *Current* | 6334 (6.9) |
| Frequency of alcohol intake | |
| *Never* | 5212 (5.7) |
| *Regularly ('1-2 times a week' /'3-4 times a week')* | 47059 (51.0) |
| *Occasionally ('1-3 times a month' / 'special occasions only')* | 18796 (20.4) |
| *Daily ('daily' / 'almost daily')* | 21058 (22.8) |
| Season at the time when accelerometer monitoring started | |
| *Spring* | 20032 (21.7) |
| *Summer* | 19663 (21.3) |
| *Autumn* | 24477 (26.5) |
| *Winter* | 28030 (30.4) |

CARE = circadian activity rhythm energy, SD = standard deviation.

**Supplementary Table 4. The intra-subject and inter-subject variability of CARE values in the adolescent and adults dataset.**

| | Df | Sum Sq | Mean Sq | F value | *P* value(>F) |
|---|---|---|---|---|---|
| **Adolescent** | | | | | |
| Subject | 999 | 2.00 | 0.002 | 3.71 | <0.0001 |
| Repetition | 1 | 0.001 | 0.001 | 1.76 | 0.18 |
| Residuals | 999 | 0.54 | 0.001 | | |
| **Adult** | | | | | |
| Subject | 999 | 2.93 | 0.003 | 2.34 | <0.0001 |
| Repetition | 1 | 0.002 | 0.002 | 1.34 | 0.25 |
| Residuals | 999 | 1.25 | 0.001 | | |

CARE = circadian activity rhythm energy, Df = degrees of freedom, Mean Sq = mean square, Sum Sq = sum of squares. Analysis of variance was performed in a subset of 1,000 individuals each from the adolescent and adult dataset who had at least six days of accelerometer data.

**Supplementary Table 5. The between-group variability of CARE values.**

| | Df | Sum Sq | Mean Sq | F value | *P* value(>F) |
|---|---|---|---|---|---|
| **Group** | 3 | 0.02 | 0.006 | 3.39 | 0.02 |
| **Residuals** | 38752 | 66.39 | 0.002 | | |

CARE = circadian activity rhythm energy, Df = degrees of freedom, Mean Sq = mean square, Mean Sq = mean square. Analysis of variance was performed in individuals with psychiatric disorders, such as bipolar affective disorder (n = 147), schizophrenia(n = 42), depression(n = 2,252), and in a control group (n = 36,315) from the adult dataset.

**Supplementary Table 6. Associations between relative amplitude and cognitive functions in the adolescent study (SCHEDULE-A).**

| Cognitive Scores | n | Coefficient | SE | *P* value |
|---|---|---|---|---|
| **BRI** | 1,703 | -7.13 | 7.65 | 0.35 |
| **MI** | 1,703 | -14.82 | 7.26 | 0.04 |
| **GEC** | 1,703 | -11.65 | 5.61 | 0.04 |

BRI = Behavioral Regulation Index, GEC = Global Executive Composite, MI = Metacognition Index, SE = standard error. Median regression models were used with adjusting age, sex, parental education level, family income, and primary caregiver. The significance level was set at $P < 0.017$.

**Supplementary Table 7. Associations between relative amplitude and cognitive functions in the adult dataset (UK Biobank).**

| Cognitive Scores | n | Coefficient/OR | SE | *P* value |
|---|---|---|---|---|
| **Processing/reaction speed** | 91,830 | - 41.88 | 5.22 | <0.0001 |
| **Reasoning ability** | 34,656 | 1.30 | 0.23 | 0.08 |
| **Short-term memory** | 77,439 | 0.95 | 0.11 | 0.64 |
| **Prospective memory** | 34,173 | 1.10 | 0.37 | 0.71 |

OR = odds ratio, SE = standard error. Models were adjusted for age, sex, ethnicity, Townsend score, and the season when the participant started wearing the accelerometer. Linear regression for processing/reaction speed, ordinal logistic regression for reasoning ability and short-term memory scores, and logistic regression for prospective memory were employed. The significance level was set at $P < 0.013$.

**Supplementary Table 8. Lead variants associated with CARE in the adult dataset (UK Biobank).**

| SNP | Chr:position | Nearest gene(s) | Alleles (E/A) | BETA | SE | *P* value |
|---|---|---|---|---|---|---|
| 3_49673081_ CCGGG_C | 3:49673081 | BSN, APEH | CCGGG/C | -0.001 | 0.0002 | $1.67 \times 10^{-9}$ |

CARE = circadian activity rhythm energy, Chr = chromosome, E/A = effect/non-effect alleles, position = base pair coordinate hg38, SE = standard error, SNP = single nucleotide polymorphism. Genetic association analysis was performed in related subjects of European ancestry using linear regression models adjusted for age, sex, 20 principal components of ancestry, genotyping array, and genetic correlation matrix. Only lead variants in each locus are shown above. Genes indicate all genes within the locus of interest.

**Supplementary Table 9. Significant variants associated with relative amplitude ($P < 5 \times 10^{-8}$) in the adult dataset (UK Biobank).**

| SNP | Chr:position | Nearest gene(s) | Alleles (E/A) | BETA | SE | *P* value |
|---|---|---|---|---|---|---|
| **rs1110666** | 22:17974954 | MICAL3 | G/T | 0.002 | 0.0004 | $2.01 \times 10^{-8}$ |
| rs9605481 | 22: 17982911 | MICAL3, MIR648 | A/G | 0.002 | 0.0004 | $2.29 \times 10^{-8}$ |
| rs12157484 | 22: 17972356 | MICAL3 | T/C | 0.002 | 0.0004 | $2.67 \times 10^{-8}$ |

Chr = chromosome, E/A = effect/non-effect alleles, position = base pair coordinate hg38, SE = standard error, SNP = single nucleotide polymorphism. Genetic association analysis was performed in related subjects of European ancestry using linear regression models adjusted for age, sex, 20 principal components of ancestry, genotyping array, and genetic correlation matrix. Lead variants in each locus are shown in bold. Genes indicate all genes within the locus of interest.

**Supplementary Table 10. Mendelian randomization analysis for CARE and cognitive functions using GWAS summary statistics in the adult dataset (UK Biobank).**

| Exposure | Outcome | Method | n SNPs | Beta | SE | *P* value |
|---|---|---|---|---|---|---|
| CARE | Reasoning ability | Weighted median | 109 | -59.91 | 1.53 | <0.0001 |
| | | MR-Lasso | 109 | -59.6 | 0.76 | <0.0001 |
| | | MBE | 109 | -60.15 | 3.96 | <0.0001 |
| | | MR-cML | 109 | -59.4 | 1.22 | <0.0001 |
| | | Inverse variance weighted | 109 | -59.6 | 0.76 | <0.0001 |
| CARE | Short-term memory | Weighted median | 109 | 7.94 | 0.53 | <0.0001 |
| | | MR-Lasso | 109 | 7.51 | 0.4 | <0.0001 |
| | | MBE | 109 | 8.03 | 1.4 | <0.0001 |
| | | MR-cML | 109 | 7.5 | 0.45 | <0.0001 |
| | | Inverse variance weighted | 109 | 7.51 | 0.4 | <0.0001 |
| CARE | Prospective memory | Weighted median | 109 | 16.85 | 1.23 | <0.0001 |
| | | MR-Lasso | 109 | 16.97 | 0.94 | <0.0001 |
| | | MBE | 109 | 16.67 | 3.12 | <0.0001 |
| | | MR-cML | 109 | 16.93 | 1.04 | <0.0001 |
| | | Inverse variance weighted | 109 | 16.97 | 0.94 | <0.0001 |

CARE = circadian activity rhythm energy, GWAS = genome-wide association study, MR = mendelian randomization, SE = standard error.

**Supplementary Table 11. Significant genes associated with circadian activity rhythm energy (CARE) detected by cross-tissue transcriptome-wide association analysis using UTMOST in 44 GTEx tissues.**

| Gene | Test score | *P* value |
|---|---|---|
| **QARS** | 14.29695 | $3.20 \times 10^{-7}$ |
| **SEMA3F** | 14.19545 | $3.10 \times 10^{-7}$ |
| **TRAIP** | 17.88998 | $7.28 \times 10^{-9}$ |
| **USP4** | 13.72578 | $6.88 \times 10^{-7}$ |
| **COL7A1** | 13.0528 | $1.42 \times 10^{-6}$ |
| **DOCK3** | 12.91689 | $1.40 \times 10^{-6}$ |
| **FAM212A** | 13.2909 | $2.24 \times 10^{-6}$ |
| **GPX1** | 12.51486 | $8.85 \times 10^{-7}$ |
| **APEH** | 17.2256 | $9.14 \times 10^{-9}$ |
| **C3orf18** | 12.95729 | $9.04 \times 10^{-7}$ |
| **IMPDH2** | 16.09161 | $7.03 \times 10^{-8}$ |
| **MAPKAPK3** | 17.61598 | $1.67 \times 10^{-8}$ |
| **MST1** | 16.40505 | $1.83 \times 10^{-8}$ |
